# Interference of urine tubular biomarker measurements by glycosuria: implications when using SGLT2 inhibitors

**DOI:** 10.1101/2025.06.15.25329638

**Authors:** Greco B Malijan, Daniel Chapman, Stewart Moffat, Rebecca J Sardell, Natalie Staplin, Martin J Landray, Colin Baigent, Michael G Shlipak, Richard Haynes, Joachim H Ix, William G Herrington, Michael Hill, Parminder K Judge

## Abstract

Sodium-glucose co-transporter 2 (SGLT2) inhibitors are recommended for use in adults with chronic kidney disease (CKD) and are widely prescribed. SGLT2 inhibition markedly increases urine glucose excretion, which could interfere with laboratory assays. We assessed whether assays for several key urine tubular biomarkers (alpha-1 microglobulin [α1M], dickkopf-3 [DKK-3], epidermal growth factor [EGF], interleukin-18 [IL-18], kidney injury molecule-1 [KIM-1], monocyte chemoattractant protein-1 [MCP-1], neutrophil gelatinase-associated lipocalin [NGAL], uromodulin [UMOD], and human cartilage glycoprotein-40 [YKL-40]) are affected by glycosuria using urine samples from participants with CKD. Each urine sample was divided into three aliquots, with one serving as control and the other two being spiked with glucose to reach effective concentrations of 28 mmol/l and 111 mmol/l. There was large positive mean bias [95% CI] observed at 28 mmol/l glucose concentration for IL-18 (0.10 [0.01, 0.23]) and YKL-40 (0.40 [0.32, 0.49]). The limits of agreement (LOA) for both biomarkers were wide, spanning >1 unit difference in log-transformed biomarker values. The rest of the biomarkers had narrow LOA. Modest negative mean bias at 28 mmol/l glucose concentration was observed for DKK-3 (−0.02 [−0.04, 0]), KIM-1 (−0.04 [−0.06, −0.02]), and UMOD (−0.08 [−0.11, −0.06]), with similar values observed at 111 mmol/l glucose concentration. There was no evidence of any bias in measurements of α1M, EGF, MCP-1, and NGAL. Glycosuria substantially interferes with IL-18 and YKL-40 measurements, without importantly affecting α1M, DKK-3, EGF, KIM-1, MCP-1, NGAL or UMOD.

## INTRODUCTION

Several urine biomarkers reflecting kidney tubular health and disease have been developed and may provide additional prognostic information alongside estimated glomerular filtration rate (eGFR) and urine albumin-to-creatinine ratio (uACR).^1,2^ Urine tubular biomarkers such as dickkopf-3 (DKK-3), interleukin-18 (IL-18), kidney injury molecule-1 (KIM-1), monocyte chemoattractant protein-1 (MCP-1), neutrophil gelatinase-associated lipocalin (NGAL), and human cartilage glycoprotein-40 (YKL-40) have been investigated in the context of acute kidney injury (AKI), kidney function decline, kidney drug monitoring, and tubulointerstitial fibrosis. Other biomarkers such as alpha-1-microglobulin (α1M), epidermal growth factor (EGF), and uromodulin (UMOD) have been studied for their proposed roles reflecting tubular reserve and function.

Sodium-glucose co-transporter 2 (SGLT2) inhibitors have been shown to reduce risk of kidney disease progression, risk of heart failure, and risk of adverse events of acute kidney injury.^3^ They are recommended for use in adults with chronic kidney disease (CKD)^4^ and widely prescribed. SGLT2 inhibition markedly increases urine glucose excretion, and although glycosuria is attenuated when eGFR is decreased, substantial glycosuria remains evident even in moderate-to-severe CKD.^5^ This is important to consider as there is experimental and clinical interest in utilizing urine kidney tubule biomarkers, but such glycosuria can interfere with laboratory assays. Previous work has illustrated how glycosuria importantly interferes the Jaffe reaction used to measure creatinine and account for urine tonicity in spot urine samples.^6^ Glucose interference has also been identified in other urine sample assays (e.g., uric acid, urea, total protein).^7^

As part of the development of urine biomarker analyses for evaluation in the EMPA-KIDNEY trial^8^, we aimed to assess whether assays for several key urine tubular biomarkers (α1M, DKK-3, EGF, IL-18, KIM-1, MCP-1, NGAL, UMOD, and YKL-40) are affected by glycosuria. We used urine samples from a cohort of adults with CKD with appropriate ethics approvals. Each urine sample was divided into three aliquots, with one serving as control and the other two being spiked with glucose to reach effective concentrations of 28 mmol/l and 111 mmol/l, which approximately correspond to the interquartile range of urine glucose concentrations among patients treated with SLGT2 inhibitors.

## RESULTS

The detailed laboratory and statistical methods are provided in the Supplementary Materials, including assay characteristics summarized in Supplementary Table S1. Urine samples from 139 participants with CKD were analyzed. At baseline, the mean±SD age of participants was 64±13 years; 37 (27%) were female, and 36 (26%) had diabetes (Supplementary Table S2). The mean±SD eGFR was 36.5±11.2 ml/min/1.73m^2^. The median [Q1,Q3] uACR was 52 [16,131] mg/mmol, with 15 (11%), 30 (22%), and 94 (68%) having uACR levels at <3, ≥3 to <30, and ≥30 mg/mmol, respectively.

Bland-Altman plot analyses showed large positive mean bias [95% confidence interval] at 28 mmol/l glucose for IL-18 (0.10 [0.01, 0.23]) and YKL-40 (0.40 [0.32, 0.49]), with comparable results at glucose concentration 111 mmol/l (Figure 1). The limits of agreement (LOA) for both biomarkers were wide, with LOA range spanning >1 unit difference in log-transformed biomarker values (before and after adding glucose). Despite wide LOAs estimated for IL-18 and YKL-40, >5% of observations fell outside the range for both glucose concentrations. The rest of the biomarkers had narrow LOAs. Modest negative mean bias at 28 mmol/l glucose concentration was observed for DKK-3 (−0.02 [−0.04, 0]), KIM-1 (−0.04 [−0.06, −0.02]), and UMOD (−0.08 [−0.11, −0.06)]. The estimated mean bias values were similar at 111 mmol/l glucose concentration for these biomarkers. There was no evidence of any bias in measurements of α1M, EGF, MCP-1, and NGAL.

**Figure 1.**
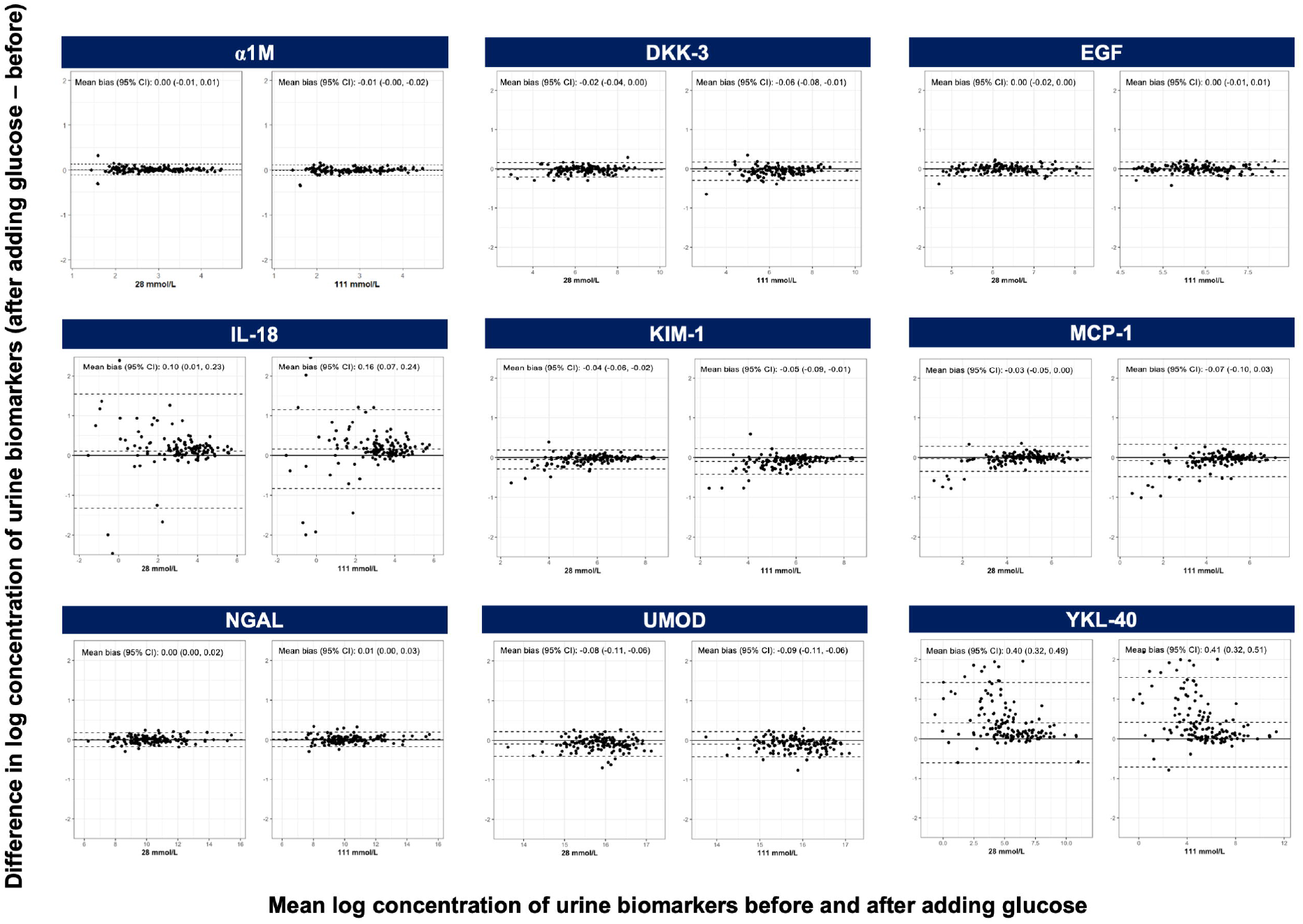
Bland-Altman plots for urine tubular biomarkers, by glucose concentration (28 mmol/l versus 111 mmol/l). Mean bias and 95% CI are shown in text. The 95% limits of agreement are shown as dashed black lines. α1M – alpha-1 microglobulin, DKK-3 – dickkopf-3, EGF – epidermal growth factor, IL-18 – interleukin-18, KIM-1 – kidney injury molecule-1, MCP-1 – monocyte chemoattractant protein-1, NGAL – neutrophil gelatinase-associated lipocalin, UMOD – uromodulin, YKL-40 – human cartilage glycoprotein-40.

## DISCUSSION

In this study evaluating the impact of glycosuria on urine tubular biomarker measurements, we found that IL-18 and YKL-40 measurements have a substantial positive bias in the presence of urine glucose, with approximately 10% and 40% overestimation respectively. Interference was evident even at lower levels of glycosuria. There was no meaningful interference for α1M, DKK-3, EGF, KIM-1, MCP-1, NGAL, and UMOD, suggesting that these biomarkers can be reliably measured in patients taking SGLT2 inhibitors using the nephelometric and electrochemiluminescence immunoassays employed in these analyses.

These findings have important implications given the expanding use of SGLT2 inhibitors in CKD management and the growing interest in urine tubular biomarkers for risk stratification and monitoring kidney health. Glucose interference likely occurs through sample matrix changes that disrupt antigen-antibody interactions and protein glycation at antigen recognition sites. Predicting which analytes are most susceptible to glycation requires detailed structural knowledge of binding sites that is not routinely available.^9^ Therefore, we recommend using glucose interference experiments as standard practice when developing urine biomarker assays in the era of SGLT2 inhibitors.

Key strengths of the study are the use of glucose concentrations reflecting the physiologic range observed with SGLT2 inhibitor therapy and evaluation of multiple biomarkers using validated assay platforms. Limitations include the *in vitro* nature of the experiments, which may not fully capture *in vivo* conditions. Moreover, we did not evaluate potential interference from other analytes that may be altered with SGLT2 inhibitor use such as ketone bodies, although such effects are unlikely to cause interference.

In summary, glycosuria substantially interferes with IL-18 and YKL-40 measurements, without importantly affecting α1M, DKK-3, EGF, KIM-1, MCP-1, NGAL or UMOD.

## Data Availability

All data produced in the present work are contained in the manuscript.

## AUTHOR’S CONTRIBUTIONS

JHI, MS, RH & WGH selected the urine biomarkers. DC, MH and SM designed and conducted the laboratory analyses, with design support provided by WGH, RH and PKJ. GBM developed and performed the statistical analyses under supervision from RS & NS. GBM, DC and SM wrote the first draft of the manuscript. All authors contributed to interpretation of the results and revision.

## RIGHTS RETENTION STATEMENT

For the purpose of open access, the authors have applied a Creative Commons Attribution (CC BY) licence to any Author Accepted manuscript version arising.

