## Supplementary Methods and Tables for "Interference of urine tubular biomarker measurements by glycosuria: implications when using SGLT2 inhibitors"

#### Supplementary Material

#### Supplementary Methods

Early morning urine samples collected and stored as part of the UK Heart and Renal Protection (HARP)-III trial (previously reported)[1] were used for laboratory interference experiments. UK HARP-III randomized 414 participants ≥18 years with CKD stages 3/4 to irbesartan or sacubitril/valsartan. The trial found that over 12 months, sacubitril/valsartan had similar effects on kidney function and albuminuria to irbesartan. Urine samples were stored long-term at -80ºC until analysis. 139 participants' samples with a urine volume of at least 1ml were used for this study.

##### Laboratory Methods

Each urine sample was thawed, mixed by inversion, and separated into three 245µl aliquots. One aliquot had 70µl of deionized water added (control sample), and the remaining aliquots were spiked with 70µl of either 22.5 or 90 g/l of glucose. The resulting spiked glucose concentrations were 28 mmol/l and 111 mmol/l, respectively. These concentrations correspond to the approximate first and third quartiles of the urine glucose levels in the empagliflozin group at 18 months in EMPA-KIDNEY, a large randomized trial of empagliflozin versus placebo among adults with CKD.[2] Each aliquot was incubated at 37°C for two hours to simulate physiologic conditions prior to analysis. Urine alpha-1 microglobulin concentrations were determined as single measurements using the Atellica Nephelometer 630 analyzer (Siemens Healthineers, Erlangen, Germany). Immunoassays for urine dickkopf-3, epidermal growth factor, interleukin-18, kidney injury molecule-1, monocyte chemoattractant protein-1, neutrophil gelatinase-associated lipocalin, uromodulin, and human cartilage glycoprotein-40 were developed in-house in multiplex panels using reagents from MesoScale Discovery on the MESO QuickPlex SQ 120 platform (MesoScale Diagnostics, Rockville, MA, USA). Sample reformatting, dilution, and electrochemiluminescence analyses were carried out using the Biomek i7-A015 Automated Liquid Handler system (Beckman Coulter, Brea, CA, USA). Multiplex panel measurements were conducted in duplicates to minimize measurement errors. All sample storage, pre-analytical preparations, assay validation, optimization, and measurements were conducted in the Wolfson Laboratory located at the University of Oxford (following the Clinical & Laboratory Standards Institute guidelines).

##### Statistical Analyses

Biomarker values were log-transformed to address skewness of biomarker concentration. Bland-Altman plots comparing the mean and difference of paired measurements of urine specimens spiked with glucose versus control were constructed using log-transformed biomarker values. Mean bias with 95% confidence interval (CI) and limits of agreement (LOA) were presented. All statistical programming were performed using R version 4.4.2 (R Foundation for Statistical Computing, Vienna, Austria) and SAS version 9.4 (SAS Institute, Cary, NC, USA).

[1] Haynes R, Judge PK, Staplin N, et al. Effects of Sacubitril/Valsartan Versus Irbesartan in Patients With Chronic Kidney Disease. *Circulation*. 2018;138(15):1505-1514. doi:10.1161/CIRCULATIONAHA.118.034818

[2] EMPA-KIDNEY Collaborative Group. Empagliflozin in Patients with Chronic Kidney Disease. *New England Journal of Medicine*. Published online November 4, 2022:1-11. doi:10.1056/NEJMoa2204233

### Supplementary Tables

#### Supplementary Table S1. Characteristics of assays used in the experiments

| Urine Assay | Dilution Factor | Concentration* | Between-plate Coefficient of Variation (%) | Limits of Detection* |
|---|---|---|---|---|
| **Nephelometry** | | | | |
| Alpha-1 microglobulin | 1 | 28.4 | 1.3 | 5.34-171 |
| **Multiplex** | | | | |
| Dickkopf-3 | 1 | 6683 | 8.6 | 10.7-44000 |
| Epidermal growth factor | 49 | 11.7 | 5.7 | 0.2-49000 |
| Interleukin-18 | 1 | 512 | 4.8 | 30-12700 |
| Kidney injury molecule-1 | 1 | 2231 | 2.6 | 19.5-20000 |
| Monocyte chemoattractant protein-1 | 1 | 624 | 4.3 | 35-6400 |
| Neutrophil gelatinase-associated lipocalin | 49 | 19300 | 14.1 | 8.4-1681680 |
| Uromodulin | 49 | 49637 | 7.1 | 1465-294000000 |
| Human cartilage glycoprotein-40 | 1 | 1418 | 7.8 | 37-18160 |

*Concentration and limits of detection reported with mg/l for alpha-1 microglobulin and pg/ml for the biomarkers measured using multiplex panels; concentration refers to the biomarker levels at which the coefficients of variation were determined

#### Supplementary Table S2. Baseline characteristics of participants with urine samples included in the laboratory interference experiment

| Characteristic | Included (N=139) |
|---|---|
| **DEMOGRAPHICS** | |
| Age at randomization, years | 64±13 |
| Female sex, no. (%) | 37 (26.6) |
| Race, no. (%)† | |
| Black | 3 (2.2) |
| South Asian | 1 (0.7) |
| Other | 5 (3.6) |
| White | 130 (94.5) |
| History of diabetes, no. (%)‡ | 36 (25.9) |
| Systolic | 146±16 |
| Diastolic BP | 82±11 |
| Body mass index, kg/m$^2$ | 30±5 |
| **LABORATORY MEASUREMENTS** | |
| Estimated glomerular filtration rate, ml/min/1.73 m$^2$ | |
| Mean ± SD | 36.5±11.2 |
| Distribution, no. (%) | |
| <30 | 47 (33.8) |
| ≥30 to <45 | 59 (42.5) |
| ≥45 | 32 (23.1) |
| Urine albumin-to-creatinine ratio, mg/mmol | |
| Median [Q1, Q3] | 52 [16, 131] |
| Distribution, no. (%) | |
| <3 | 15 (10.8) |
| 3 to 30 | 30 (21.6) |
| >30 | 94 (67.6) |
| RAS inhibitor use at baseline, no. (%) | 116 (83.5) |

* Plus-minus values are means ± SD. 2 participants had missing body mass index measurements, and 1 participant had missing estimated glomerular filtration rate at baseline.
† Race was reported by the patients. The “other” category indicates that the race was not specified or the patient preferred not to answer.
‡ History of diabetes was defined as patient-reported history of diabetes of any type, use of glucose-lowering medication, or a glycated haemoglobin level of at least 48 mmol/mol at the randomization visit.
